## Supplementary Material for "DNA mismatch repair gene variant classification: evaluating the utility of somatic mutations and mismatch repair deficient colonic crypts and endometrial glands"

Department of Clinical Pathology

The University of Melbourne

Victorian Comprehensive Cancer Centre

305 Grattan Street

Parkville, Victoria, 3010 Australia

Ph: +61 0401562006

### SUPPLEMENTARY FIGURES

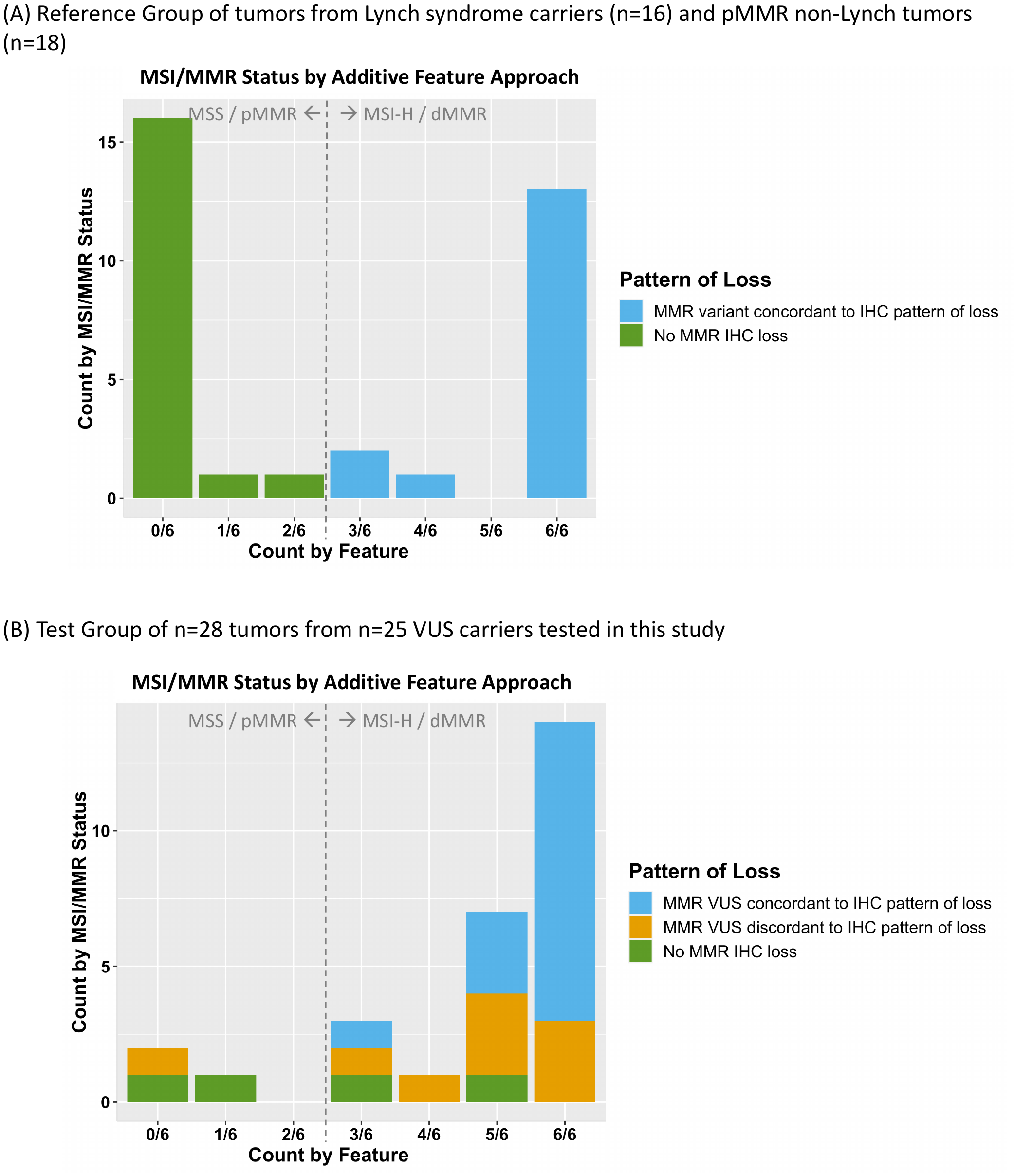

##### **Figure S1.** Bar plot presenting the feature count for targeted panel sequenced (A) reference and (B) test groups to determine the DNA mismatch repair status in next-generation sequencing screened tumors after applying the additive feature combination approach. Grey dotted line indicates MSI and MMR status by additive feature combination approach with 0/6, 1/6 and 2/6 indicating MSS/pMMR and 3/6, 4/6, 5/6 and 6/6 indicating MSI-H/dMMR. *Abbreviations:* MMR, DNA mismatch repair; dMMR, DNA mismatch repair deficient; pMMR, DNA mismatch repair proficient; MSI, microsatellite instability; MSI-H, high levels of microsatellite instability; MSS, microsatellite stable; IHC, immunohistochemistry; VUS, variant of uncertain clinical significance.

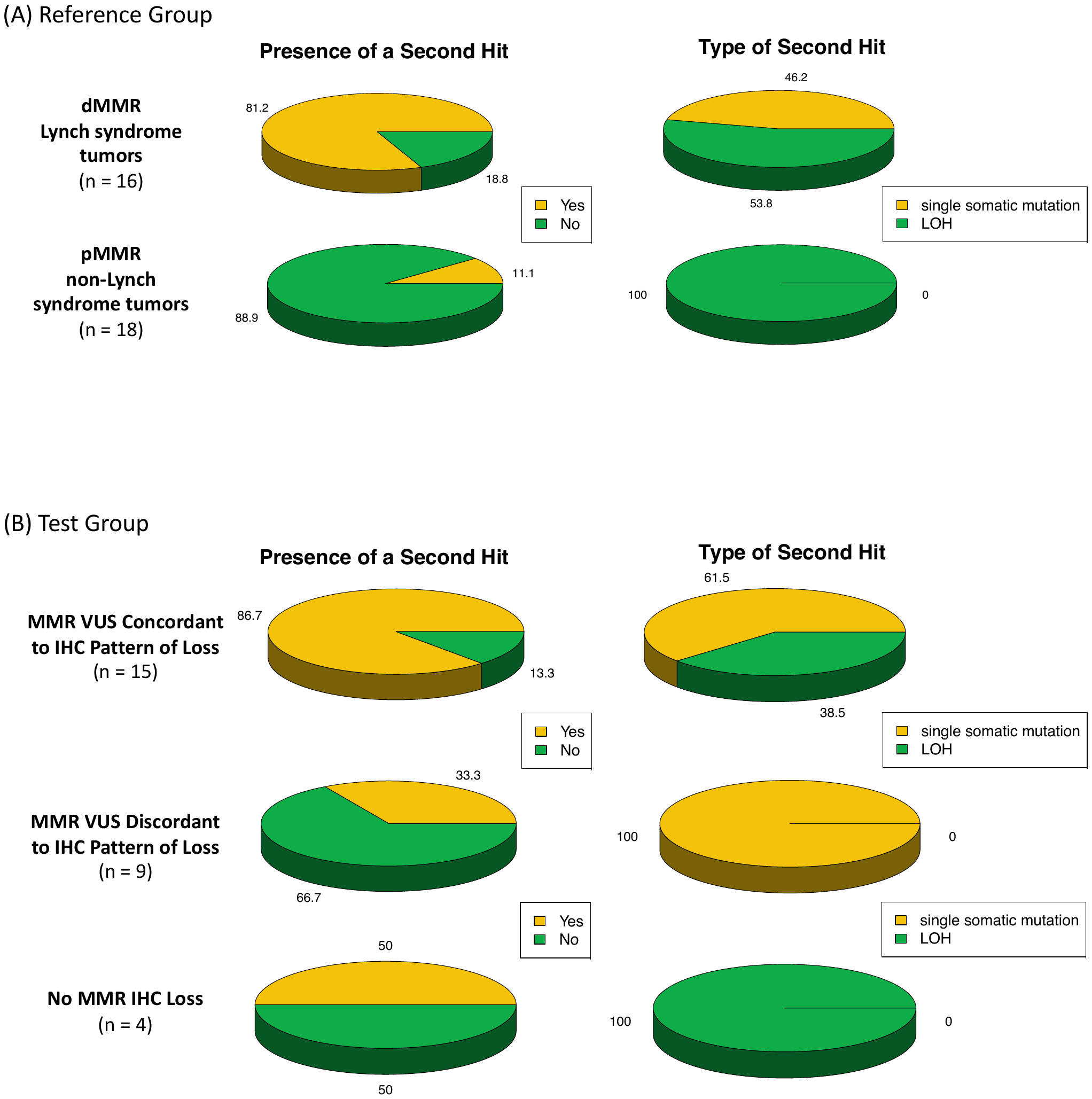

##### **Figure S2.** Pie charts presenting the proportions of the presence of a somatic second hit and by which mutation type for (A) the reference and (B) the test groups. Abbreviations: MMR, DNA mismatch repair; dMMR, DNA mismatch repair deficiency, pMMR, DNA mismatch repair proficiency; VUS, variant of uncertain significance; IHC, immunohistochemistry; LOH, loss of heterozygosity.

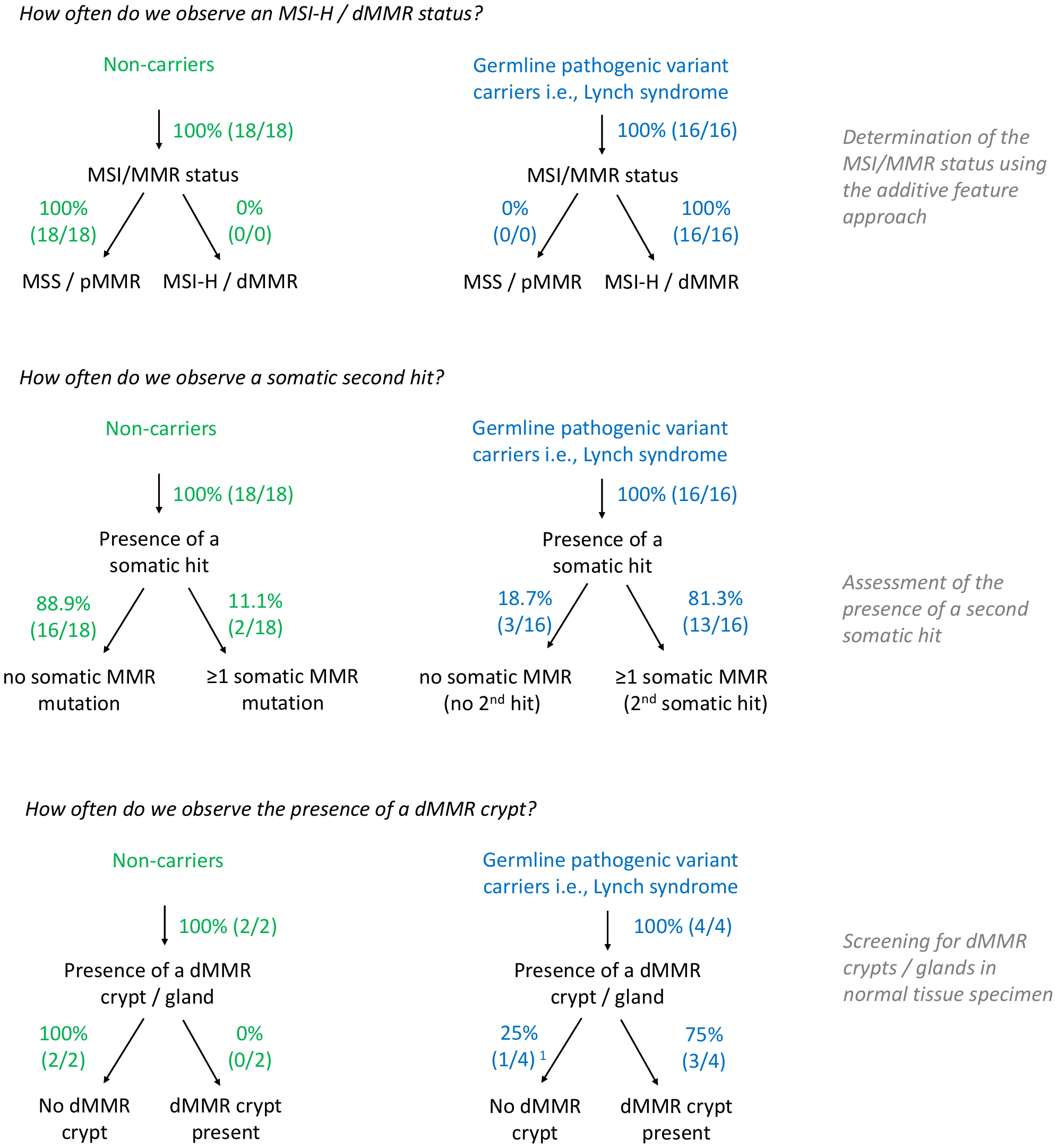

##### **Figure S3.** Flowchart displaying the prevalence of the three Lynch syndrome associated features in the reference group. Flowchart incorporating the tumor sequencing and dMMR crypt/gland screening for determining pathogenicity against the ACMG/InSiGHT framework for MMR VUS in the reference group. *Abbreviations*: MMR, DNA mismatch repair; dMMR, DNA mismatch repair deficient; pMMR, DNA mismatch repair proficient; MSI, microsatellite instability; MSI-H, high levels of microsatellite instability; MSS, microsatellite stable; VUS, variant of uncertain clinical significance; NGS, next-generation sequencing; LP, likely pathogenic.

##### ^1^ Block was depleted after screening of 2x80µM of normal colorectal tissue.

### SUPPLEMENTARY TABLES

##### **Supplementary Table 1.** Overview of tumors included in the reference group.

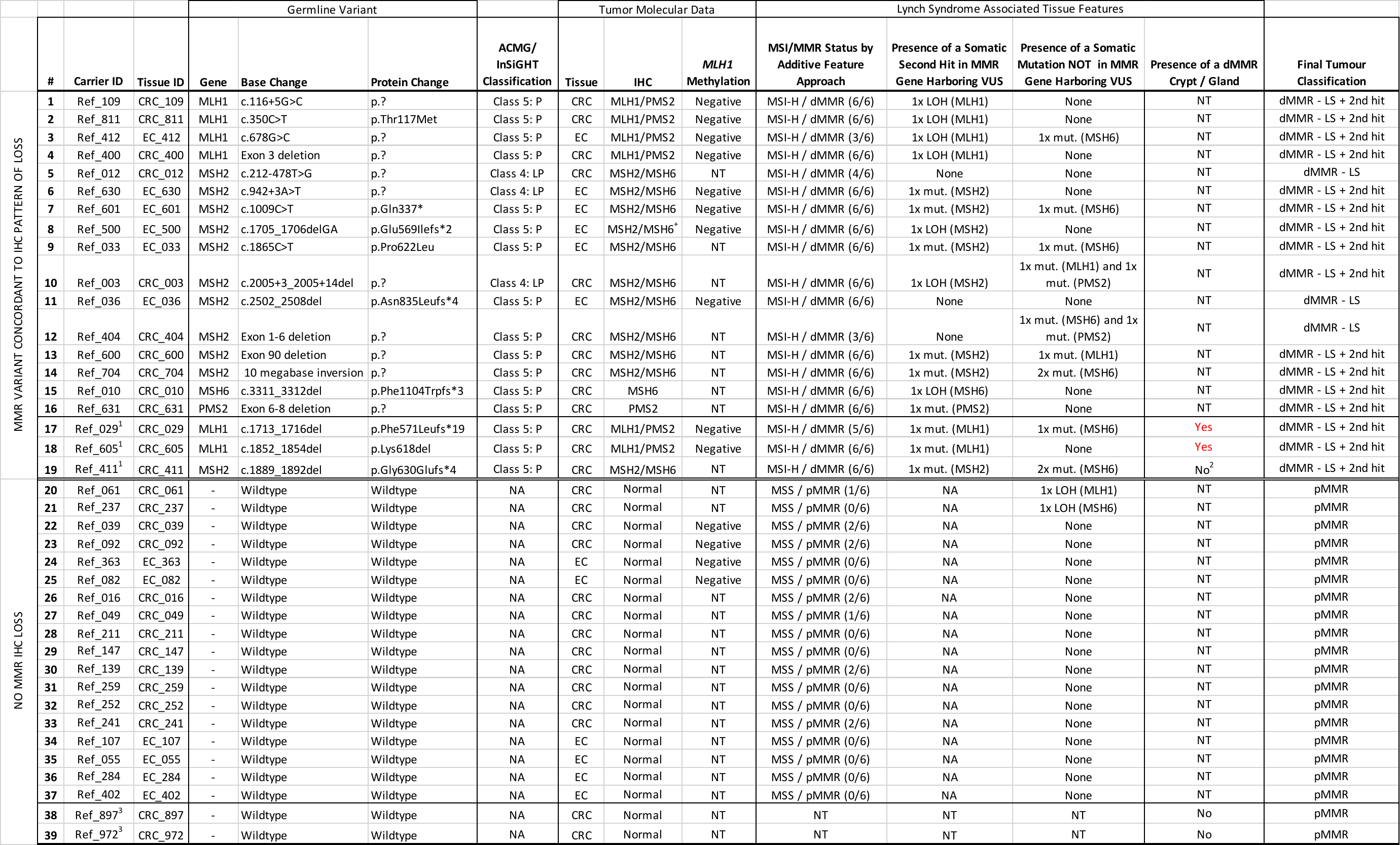

*Abbreviations*: ID, identification number; CRC, colorectal cancer; EC, endometrial cancer; IHC, immunohistochemistry; P, pathogenic; LP, likely pathogenic; MMR, DNA mismatch repair; MSI-H, high levels of microsatellite stability; MSS, microsatellite stable; dMMR, DNA mismatch repair deficient; pMMR, DNA mismatch repair proficient; LOH, loss of heterozygosity; mut., single somatic mutation; NA, not applicable; NT, not tested; LS, Lynch syndrome.

^+^ Indicates heterogeneous / patchy loss of DNA mismatch repair protein expression by IHC

^1^ These samples have undergone whole-exome sequencing

^2^ Block was depleted after screening of 2x80µM of normal tissue

^3^ These samples did not undergo next-generation sequencing
